# Decreased cerebral blood flow in a supine position in patients with a history of syncope and presyncope during the Tilt Table Test

**DOI:** 10.1101/2024.03.21.24303881

**Authors:** V Cano, JA González-Hermosillo, Ch Murata, M Martínez-Rosas, G Hernández-Pacheco

**Affiliations:** Departamento de Fisiología, Instituto Nacional de Cardiología Ignacio Chávez. Ciudad de México, México; Clínica de disautonomía, Instituto Nacional de Cardiología Ignacio Chávez. Ciudad de México, México; Departmento de Metodología de la Investigación, Instituto Nacional de Pediatría. Ciudad de México, México

**Keywords:** Presyncope, Cerebral blood flow, Transcranial Doppler, Hemodynamic response, Tilt-table test

## Abstract

**Objective:** To analyze the hemodynamic response, cerebral blood flow, and the sympathovagal balance between patients with structurally healthy hearts who have a history of syncope and presyncope that exhibited signs or symptoms of presyncope without progressing to syncope during the tilt-table test, and a healthy group.

**Methodology:** A retrospective study was conducted on patients attending the dysautonomia clinic of a tertiary care center. Individuals of both sexes aged 15 to 50 years were included. Patients with positive tests for VVS, OH, or POTS, and those with negative results were excluded. As a healthy group, healthy volunteers without a personal history of syncope or presyncope and with a negative tilt-table test were included.

**Statistical Analysis:** Categorical variables were expressed as percentages and quantitative variables as mean ± standard deviation. Repeated measures ANOVA, the χ2 test, and effect size using Glass’s Δ were used for group comparisons. A P-value < 0.05 was considered statistically significant. The R software, RStudio, and the effsize package were used for analysis.

**Results:** The main finding was a significant decrease in cerebral blood flow secondary to an increase in cerebral vascular resistance in patients from a supine position at the start and throughout the test, compared to the control group.

**Conclusion:** The analysis of cerebral hemodynamics allows for the identification of a subpopulation of patients who exhibit decreased cerebral blood flow from a supine position during the tilt-table test while maintaining cardiovascular stability without progressing to syncope.

## Introduction

Acute cerebral hypoperfusion can be a cause of syncope and presyncope symptomatology. Syncope is defined as a sudden and transient loss of consciousness, characterized by a rapid onset, short duration, and spontaneous recovery (1,2,3), while presyncope is generally defined as a sudden sensation of impending loss of consciousness with characteristic symptoms and signs such as dizziness, nausea, sensations of heat or cold, abdominal pain, visual disturbances, pallor, diaphoresis, trembling, sighs, confusion, and feeling of vomiting (4). The brain is a specialized organ with a high metabolic rate and limited energy reserves, making it essential that its blood flow is maintained at optimal levels to ensure function. From a hemodynamic perspective, cerebral blood flow (CBF) depends on mean arterial pressure, intracranial pressure, and cerebrovascular resistance, and its regulation is highly controlled (5). Syncope and presyncope are common reasons for visits to emergency services (4,6), with a reported prevalence that varies depending on the studied group. For example, in military personnel, the incidence of syncope has been estimated at 7.2 cases per 1000 persons per year, and in operating room staff, 4.7% have experienced at least one episode of syncope, and 14.8% have experienced presyncope (7,8). One of the tests used for the diagnosis and management of syncope is the tilt-table test (TTT), a technique that allows for the reproduction of signs and symptoms, and observation of the heart rate (HR) and blood pressure (BP) response to controlled postural changes (9,10). This has led to the identification of various conditions such as orthostatic hypotension (OH), postural orthostatic tachycardia syndrome (POTS), vasovagal syncope (VVS), reflex syncope (RS), among others (3). During the tilt-table test, we have observed patients who exhibit presyncope signs and symptoms without progressing to full syncope, and without showing a cardiovascular response indicative of conditions such as VVS, SN, POTS, or OH. We hypothesize that these patients have an alteration in the regulation of CBF that leads them to present symptoms without reaching syncope. Therefore, the objective of this study was to compare the CBF, cardiovascular response, and sympathovagal balance during the TTT between these types of patients and a control group.

## Materials and Methods

A retrospective study was conducted on patients who presented at the dysautonomia clinic in a tertiary level center. Prior to the study, informed consent was obtained, and for minors under 18 years, informed assent was obtained, and the responsible caregiver signed the consent. The protocol was approved by the Ethics and Research Committees of the National Institute of Cardiology Ignacio Chavez (14-904).

### Participants

The patient database with a history of syncope and/or presyncope who underwent a TTT at the dysautonomia clinic of the Institute from January 2012 to December 2016 was reviewed. Individuals of both sexes aged 15 to 50 years, with a non-pharmacological challenge test and measurement of CBF velocity, without cardiovascular disease, hypertension, diabetes mellitus, cancer, or known neurological diseases were included. Those who presented signs and/or symptoms of presyncope, reaching the end of the test without syncope, were included. Patients with a negative test, those with positive tests for POTS, OH, VVS were excluded. Tests with incomplete data or artifacts in the record were removed.

In the healthy group, healthy volunteers, medical students, and institute staff, without a personal history of syncope or presyncope and with a negative test, were included. Positive tests and tests with incomplete data or artifacts in the record were excluded.

### Tilt-Table Test

All participants underwent the tilt-table test under conditions of overnight fasting, without having consumed caffeinated beverages, alcohol, or smoked in the 48 hours prior to the test, in a semi-dark, quiet room at a temperature of 23 to 24 °C. For women of childbearing age, the test was carried out during the proliferative phase of the menstrual cycle.

In preparing the participants, the subject was placed in a supine position on the tilt-table, electrodes were attached for continuous electrocardiogram (ECG) recording; a digital plethysmograph on the second phalanx of the left index or middle finger for continuous BP measurement, an automatic oscillometer on the right arm to periodically calibrate the digital finger signal, all connected to the TaskForce Monitor (TFM, CNSystems, Graz Austria). CBF velocity was measured using transcranial Doppler ultrasound (MultiDop T; DLW, Electronics, Sippligen, Germany) coupled to the TaskForce Monitor, a transducer was placed in the left parietal region (middle cerebral artery) of the participant and fixed to the skull with a band, the Doppler. A saline solution venoclysis was placed to maintain a permeable pathway, and the subject was kept in a supine position for 10 minutes for vital sign stabilization.

The test was carried out in three stages, in the following order: basal stage in supine position for five minutes, transition phase from supine to standing position, orthostatic stage at 70° incline for 30 minutes followed by the recovery stage for five minutes back in the supine position. The transition phase was excluded from the analysis.

### Definitions According to Cardiovascular Response in the TTT

Syncope: Decrease in HR ≤40bpm and/or fall in systolic BP ≤60 mmHg (2,10).

Orthostatic Hypotension: progressive and sustained decrease in systolic BP ≥20mm Hg or ≥10 mm Hg within the first minutes of orthostatism (2).

Postural Orthostatic Tachycardia Syndrome: orthostatic HR increase (>30bpm or >120 bpm within 10 minutes of active standing) in the absence of OH that reproduces spontaneous symptoms (2). Positive Test: when the subject presents a decrease in BP and/or HR in addition to signs and symptoms indicating the imminence of syncope.

Negative Test: absence of syncope and no decrease in BP and/or HR plus the absence of signs or symptoms.

Patients: when the patient did not present syncope or alterations in BP and/or HR that allowed the diagnosis of VVS, OH, POTS, or others, and presented signs and/or symptoms such as dizziness, nausea, headache, sensations of heat or cold, abdominal pain, visual disturbances, pallor, diaphoresis, trembling, confusion, feeling of vomiting, etc.

### Data Collection

The variable recording was carried out beat-to-beat, from the TFM device the average of each variable was acquired every five minutes throughout the test. The cardiovascular response variables were: HR [bpm], mean arterial pressure (MAP) [mmHg] and total peripheral resistance index according to the subject’s body surface area (TPRI) [dyns*s*m^2^/cm^5^]; from the cerebral vascular response, CBF [cm/s] and cerebral vascular resistance (CVR) [mmHg*s/cm] were measured; and from spectral analysis, the sympathovagal balance (LF/HF) of the low (LF) and high frequency (HF) components was obtained.

### Statistical Analysis

Demographic and physical characteristics of the participants were described in each group, categorical variables were expressed as a percentage, quantitative variables as mean ± standard deviation or as median 1st and 3rd quartile. The comparison between groups was analyzed by the technique of repeated measures ANOVA, also determining the difference between groups every 5 minutes throughout the test. The difference in sex distribution between the two groups was determined by the χ2 test. Due to the lack of homoscedasticity in several variables, the Welch test was used in the comparison of variables. The effect size was estimated by Glass’s Δ to not assume equality of variance between the two groups. With the sex variable, the effect size was expressed by the φ index. Point estimates of the effect size were reported with a 95% confidence interval. As a method of controlling the problem of type I error inflation, the Benjamini-Hochberg method (11) was used. Differences that were significant after correcting by this method were indicated with asterisks: P < 0.05 with “*” and P < 0.01 with “**”. Statistical analyses were performed using R (R Core Team, 2016) and RStudio (RStudio Team, 2015), as well as the “effsize” package (Torchiano, 2016).

Independent data access and analysis: In compliance with the AHA Journals Research Guidelines, we confirm that Dr. Guadalupe Hernandez Pacheco, PhD, has had full access to all the data in the study and takes full responsibility for the integrity of the data and the accuracy of the data analysis. The Editors reserve the right to request additional information from the corresponding author regarding measures taken to minimize bias and verify the integrity of the primary data and any analyses performed.

## Results

This section presents the outcomes of a comparative analysis between a group of 32 patients and a group of 40 healthy. Figure 1 displays the flowchart for selecting the study groups and Table 1 outlines their demographic characteristics. It was found that the weight and body mass index (BMI) of the patients were significantly lower compared to the healthy group (P=0.009, P=0.0026 respectively); no other characteristic differences were observed. During the orthostatic stage of the test, patients exhibited one or more of the following signs and symptoms: dizziness (40.6%), headache (34.4%), nausea (28.1%), hyperventilation (18.8%), undefined discomfort (17.5%), weakness (12.5%), crying (12.5%), diaphoresis (12.5%), among others.

**Figure 1.**
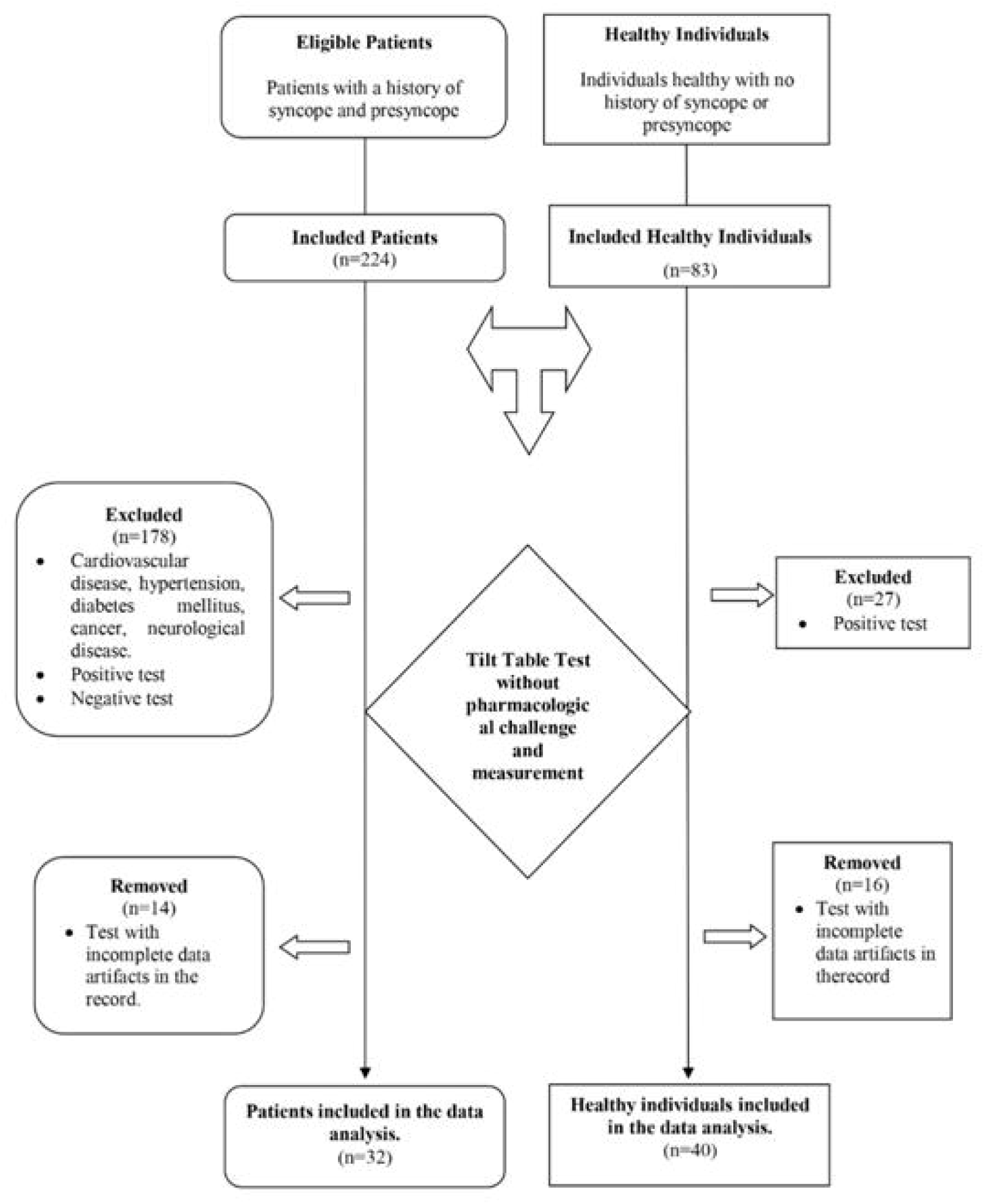
Flow chart for participants included in the study.

**Table 1.** Demographic and physical characteristics of patients and healthy subjects.

| Variables | Group |  | ES<br>[95%CI] | *p* |
| --- | --- | --- | --- | --- |
|  | Patients<br>(n=32) | Healthy<br>(n=40) |  |  |
| Age [years] | 28.4 (9.1) | 26.3 (5.6) | 0.38 [-0.09, 0.86] | 0.2510 |
| Sex [Female] | 20 (63%) | 17 (43%) | -0.02 [-0.50, 0.45] | 0.0916 |
| Height [m] | 1.68 (0.10) | 1.70 (0.11) | -0.16 [-0.63, 0.32] | 0.4898 |
| Body weight [kg] | 64.5 (11.2) | 72.8 (15.1) | -0.55 [-1.03, -0.07] | **0.0090** |
| BMI [kg/m<sup>2</sup>] | 22.7 (2.7) | 25.1 (3.8) | -0.63 [-1.12, -0.15] | **0.0026** |

ES: Effect Size (Glass's). 95%CI: 95%Confidence Interval .*p*: P-value from Welch's test.

Figure 2 shows the graphs describing the comparative cardiovascular response between groups. An increase in heart rate (HR) was observed, and while it was higher in patients, it was not significantly different from the healthy group. Mean arterial pressure (MAP) of patients in basal position was lower than control; it subsequently remained at similar values between both groups throughout the test, while the response of the total peripheral resistances was the same in both groups. Figure 3 shows the graphs describing the comparative cerebral hemodynamics between groups, the graph shows a significant difference in the decrease (p<0.0001) of CBF in patients from the supine position, during the orthostatic stage and was maintained in the recovery stage compared to the control group. Likewise, a significant increase in cerebral vascular resistance (CVR) was observed from the supine position to recovery stage in patients compared to the control group (p<0.0001). Regarding autonomic response, the change towards sympathetic tone increased significantly in the patients from minute 15 (p=0.0232) of the test and remained higher compared to the healthy group even in recovery (p<0.01).

**Figure 2.**
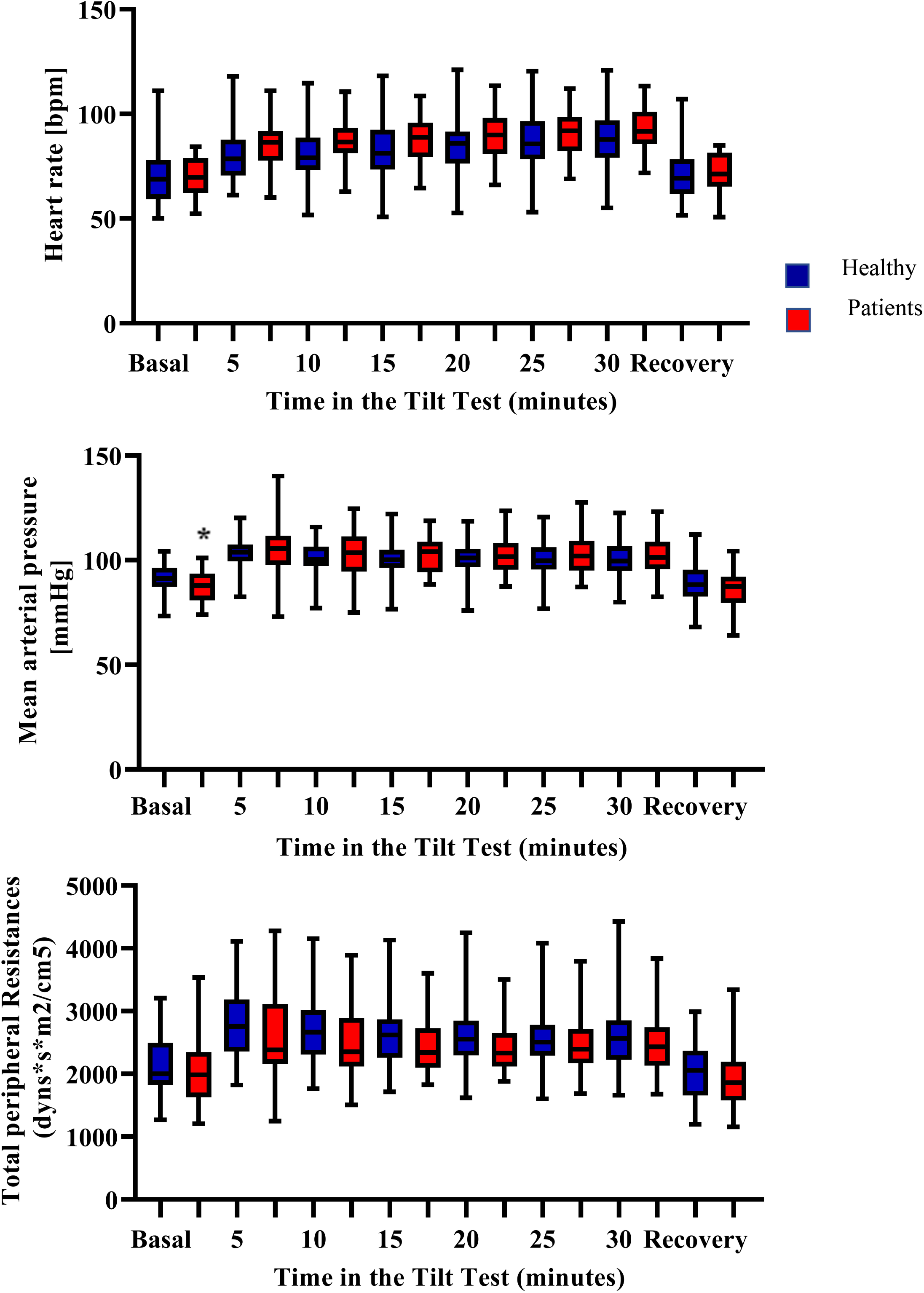
Graph of the comparison in the response of heart rate, mean arterial pressure and total peripheral resistances between healthy individuals, and patients during tilt table test. Asterisk indicates a significant difference between groups (p-value < 0.05).

**Figure 3.**
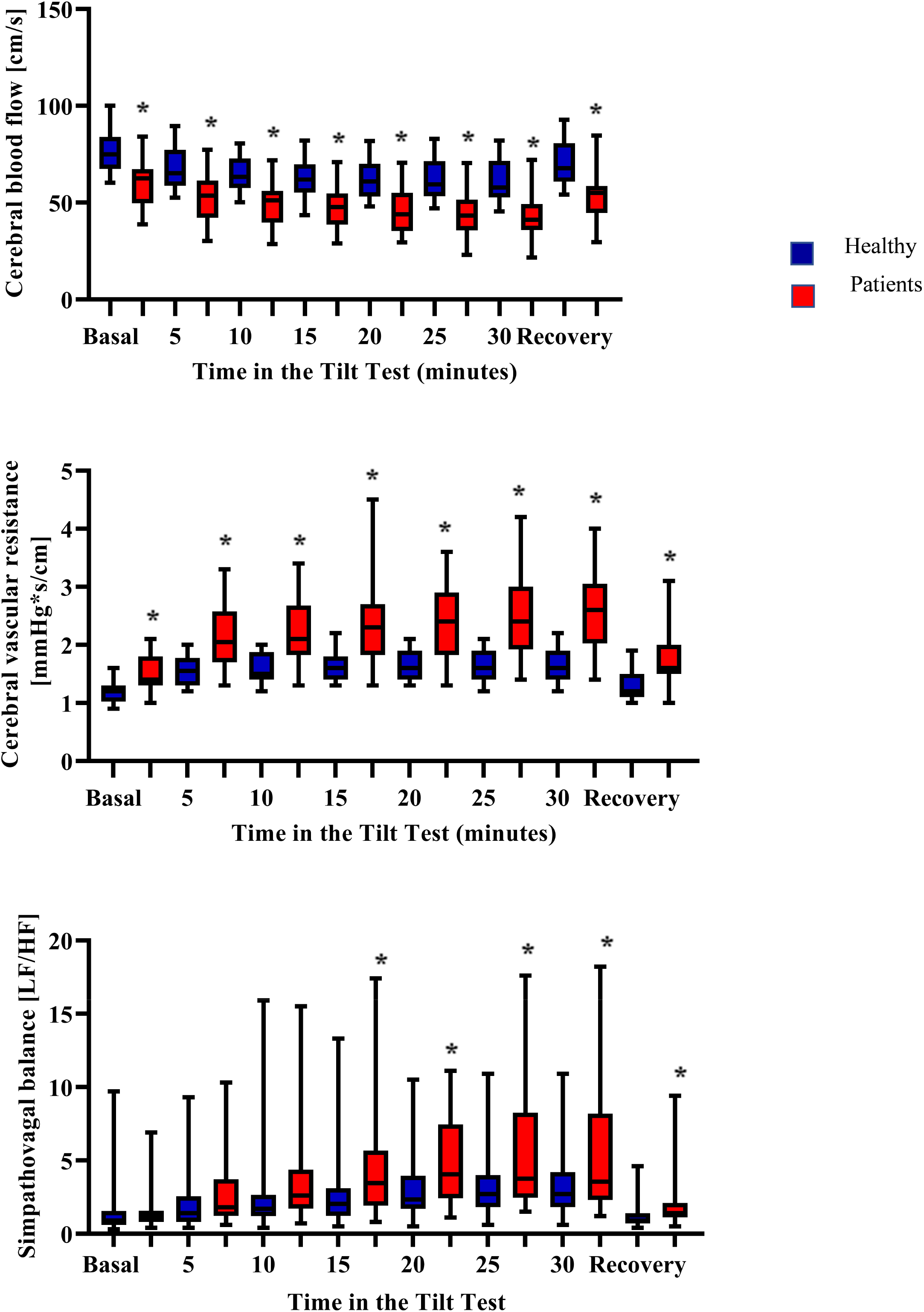
Graph of the comparison in the response of cerebral blood flow, cerebral vascular resistance and sympathovagal balance between healthy individuals and patients during tilt table test. Asterisk indicates a significant difference between groups (p-value < 0.05).

Table 2 presents the results of the analysis of variance of the response variables in the orthostatic maneuver of the groups. There was a main effect of time in the test (p<0.001), with no significant interaction between the GroupTime factors in HR, MAP, and TPR. There was a main effect of the Group and Time factor (p<0.001 each) in CBF and CVR, with significant interaction in the GroupTime factors in the CVR analysis; whereas in the analysis of the sympathetic-vagal balance, there was a main effect of the Group factor (p=0.006) and time (p<0.001) and significant interaction of Group*Time (p=0.013).

**Table 2.**
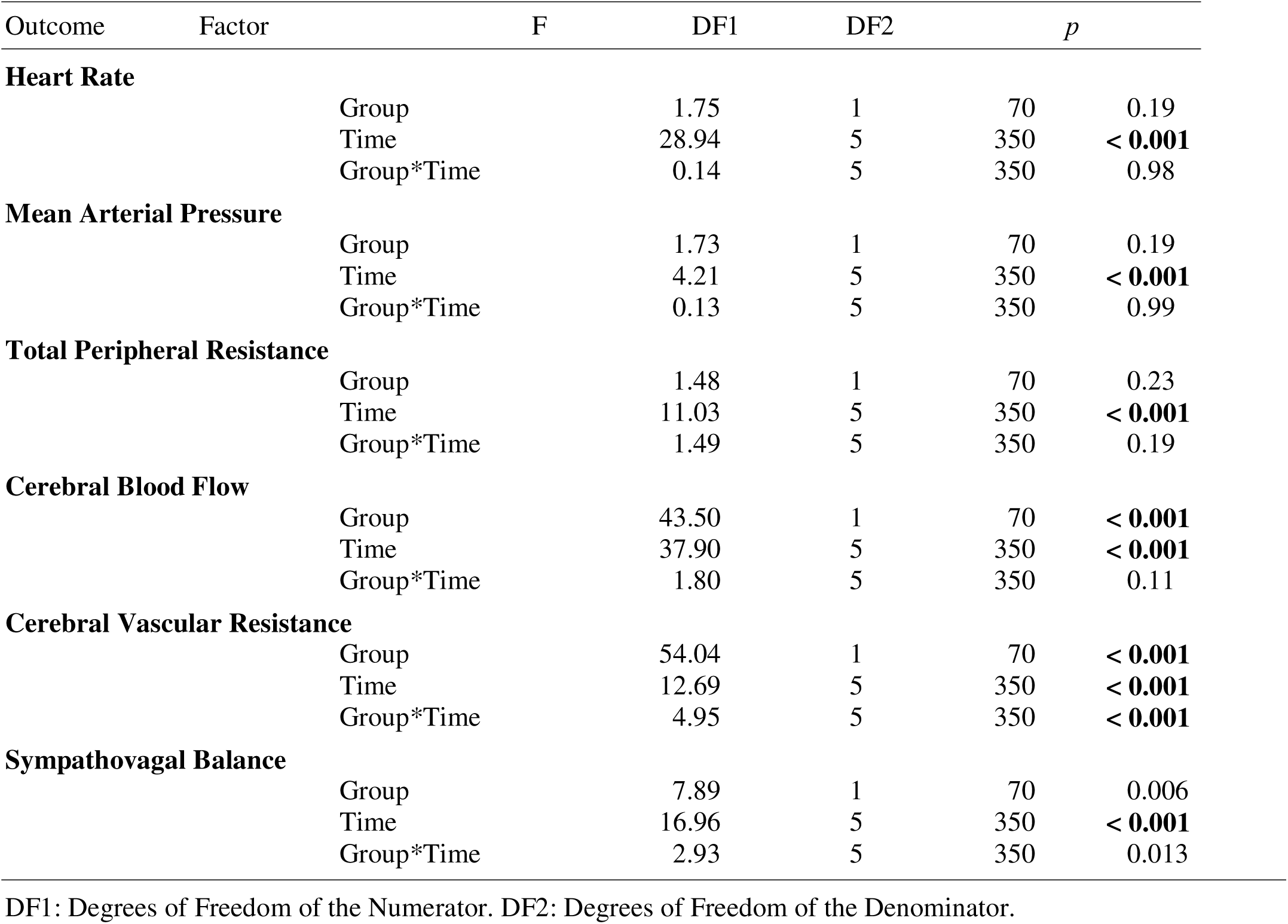
Analysis of variance of repeated measures analysis if the parameters measured during the test.

Table 3 shows the results of the comparison of cardiovascular and cerebral hemodynamic parameters between groups during the test. There was no difference between groups in heart rate during the three stages of the test, with a small effect size. The MAP values in the patients were lower in the baseline stage compared to the healthy group (p=0.0412), but this difference was lost in the subsequent stages. The mean values of the TPR in both groups were similar throughout the test.

**Table 3.** Comparisonof cardiovascular parameters between patients and healthy subjects during the test.

| Parameter | Group |  | ES<br>[95%CI] | p |
| --- | --- | --- | --- | --- |
|  | Patients<br>(n=32) | Healthy<br>(40) |  |  |
| **Hearth Rate** |  |  |  |  |
| Baseline | 69.9 (9.3) | 70.2 (13.1) | -0.02 [-0.50, 0.45] | 0.9081 |
| 5 min | 85.0 (11.6) | 80.7 (13.8) | 0.32 [-0.16, 0.79] | 0.1482 |
| 10 min | 86.7 (11.4) | 82.6 (14.8) | 0.28 [-0.20, 0.75] | 0.1909 |
| 15 min | 88.2 (11.2) | 83.9 (14.7) | 0.29 [-0.19, 0.76] | 0.1678 |
| 20 min | 89.6 (11.5) | 85.9 (14.7) | 0.25 [-0.22, 0.73] | 0.2362 |
| 25 min | 90.9 (11.2) | 87.2 (14.0) | 0.26 [-0.21, 0.74] | 0.2185 |
| 30 min | 91.9 (10.7) | 88.6 (14.2) | 0.24 [-0.24, 0.71] | 0.2575 |
| Recovery | 71.8 (9.4) | 71.4 (12.1) | 0.03 [-0.44, 0.50] | 0.8808 |
| **Mean Arterial Pressure** |  |  |  |  |
| Baseline | 87.9 (7.7) | 91.6 (6.9) | -0.53 [-1.0, -0.05] | **0.0412\*** |
| 5 min | 105.4 (12.0) | 103.2 (8.4) | 0.27 [-0.21, 0.74] | 0.3704 |
| 10 min | 102.8 (11.1) | 100.6 (8.3) | 0.27 [-0.21, 0.74] | 0.3563 |
| 15 min | 102.2 (8.8) | 99.5 (8.8) | 0.31 [-0.17, 0.78] | 0.1992 |
| 20 min | 102.2 (9.2) | 100.1 (8.6) | 0.25 [-0.23, 0.72] | 0.3168 |
| 25 min | 103.4 (9.7) | 100.1 (8.7) | 0.38 [-0.10, 0.85] | 0.1398 |
| 30 min | 103.0 (9.3) | 100.4 (9.2) | 0.28 [-0.20, 0.75] | 0.2466 |
| Recovery | 86.2 (6.6) | 88.9 (10.3) | -0.26 [-0.74, 0.21] | 0.2284 |
| **Total Peripheral Resistances** |  |  |  |  |
| Baseline | 2,156 (487) | 2,028 (567) | -0.26 [-0.74, 0.21] | 0.3153 |
| 5 min | 2,810 (572) | 2,605 (699) | -0.36 [-0.84, 0.12] | 0.1846 |
| 10 min | 2,709 (571) | 2,523 (569) | -0.33 [-0.80, 0.15] | 0.1729 |
| 15 min | 2,641 (521) | 2,473 (491) | -0.32 [-0.79, 0.16] | 0.1685 |
| 20 min | 2,612 (529) | 2,446 (440) | -0.31 [-0.79, 0.16] | 0.1505 |
| 25 min | 2,573 (506) | 2,496 (486) | -0.15 [-0.63, 0.32] | 0.5151 |
| 30 min | 2,580 (543) | 2,494 (499) | -0.16 [-0.63, 0.32] | 0.4873 |
| Recovery | 2,044 (454) | 1,936 (536) | -0.24 [-0.71, 0.24] | 0.3679 |

ES: Effect Size (Glass's $\Delta$ ). 95%CI: Confidence Interval 95%. p: P-value from Welch's test. min: minutes. The value for "Baseline" was measured at 0 minutes and "Recovery" at 35 minutes. Glass's $\Delta$ was calculated with the patient group as the reference group.
\*Statistically significant at $P < 0.05$ level with Benjamini-Hochberg correction

**Table 4.**
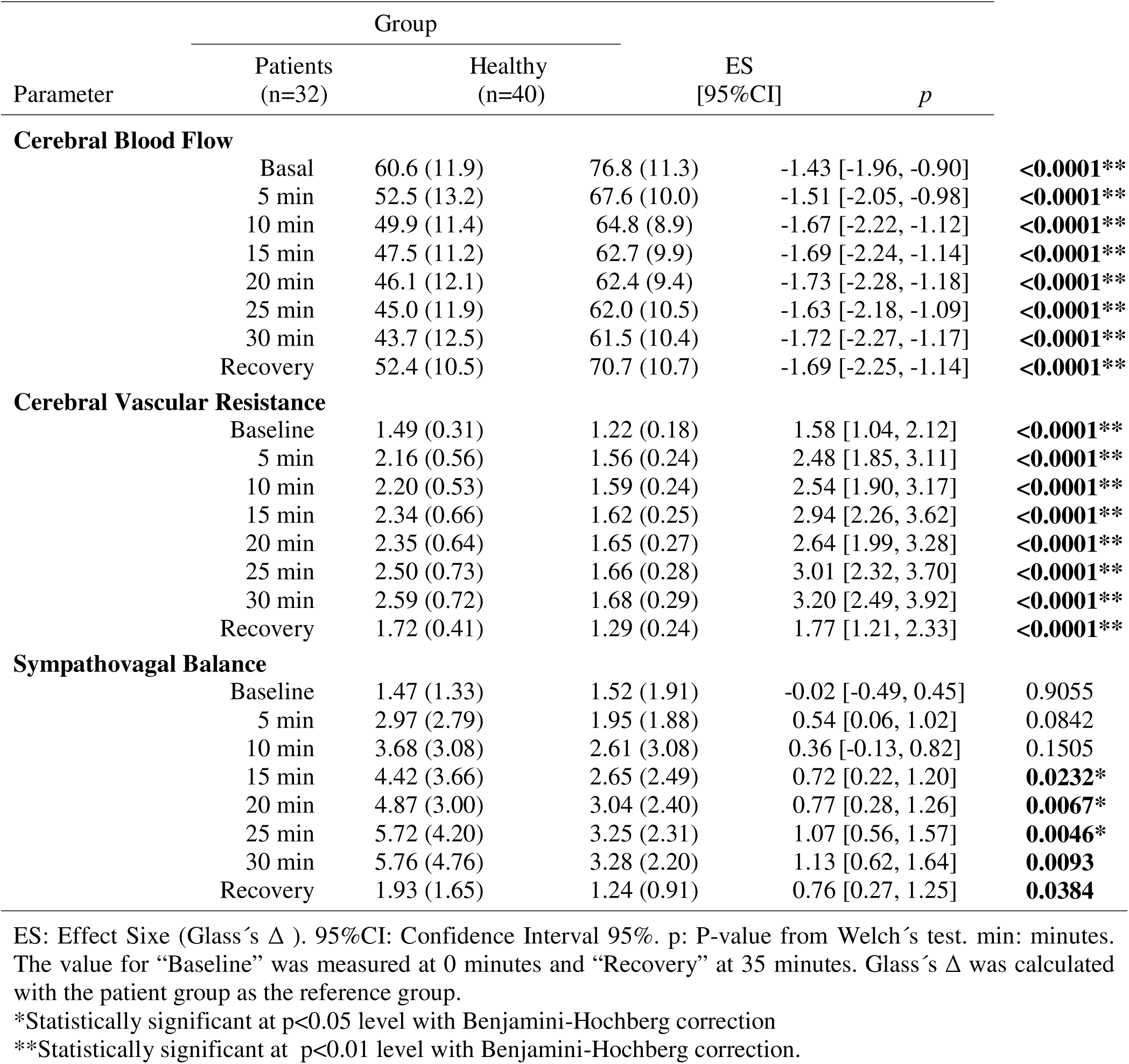
Comparison of cerebrovascular parameters and sympathovagal balance between patients and healthy subjects during the test.

| Parameter | Group |  | ES<br>[95% CI] | p |
| --- | --- | --- | --- | --- |
|  | Patients<br>(n=32) | Healthy<br>(n=40) |  |  |
| **Cerebral Blood Flow** |  |  |  |  |
| Basal | 60.6 (11.9) | 76.8 (11.3) | -1.43 [-1.96, -0.90] | **<0.0001\*\*** |
| 5 min | 52.5 (13.2) | 67.6 (10.0) | -1.51 [-2.05, -0.98] | **<0.0001\*\*** |
| 10 min | 49.9 (11.4) | 64.8 (8.9) | -1.67 [-2.22, -1.12] | **<0.0001\*\*** |
| 15 min | 47.5 (11.2) | 62.7 (9.9) | -1.69 [-2.24, -1.14] | **<0.0001\*\*** |
| 20 min | 46.1 (12.1) | 62.4 (9.4) | -1.73 [-2.28, -1.18] | **<0.0001\*\*** |
| 25 min | 45.0 (11.9) | 62.0 (10.5) | -1.63 [-2.18, -1.09] | **<0.0001\*\*** |
| 30 min | 43.7 (12.5) | 61.5 (10.4) | -1.72 [-2.27, -1.17] | **<0.0001\*\*** |
| Recovery | 52.4 (10.5) | 70.7 (10.7) | -1.69 [-2.25, -1.14] | **<0.0001\*\*** |
| **Cerebral Vascular Resistance** |  |  |  |  |
| Baseline | 1.49 (0.31) | 1.22 (0.18) | 1.58 [1.04, 2.12] | **<0.0001\*\*** |
| 5 min | 2.16 (0.56) | 1.56 (0.24) | 2.48 [1.85, 3.11] | **<0.0001\*\*** |
| 10 min | 2.20 (0.53) | 1.59 (0.24) | 2.54 [1.90, 3.17] | **<0.0001\*\*** |
| 15 min | 2.34 (0.66) | 1.62 (0.25) | 2.94 [2.26, 3.62] | **<0.0001\*\*** |
| 20 min | 2.35 (0.64) | 1.65 (0.27) | 2.64 [1.99, 3.28] | **<0.0001\*\*** |
| 25 min | 2.50 (0.73) | 1.66 (0.28) | 3.01 [2.32, 3.70] | **<0.0001\*\*** |
| 30 min | 2.59 (0.72) | 1.68 (0.29) | 3.20 [2.49, 3.92] | **<0.0001\*\*** |
| Recovery | 1.72 (0.41) | 1.29 (0.24) | 1.77 [1.21, 2.33] | **<0.0001\*\*** |
| **Sympathovagal Balance** |  |  |  |  |
| Baseline | 1.47 (1.33) | 1.52 (1.91) | -0.02 [-0.49, 0.45] | 0.9055 |
| 5 min | 2.97 (2.79) | 1.95 (1.88) | 0.54 [0.06, 1.02] | 0.0842 |
| 10 min | 3.68 (3.08) | 2.61 (3.08) | 0.36 [-0.13, 0.82] | 0.1505 |
| 15 min | 4.42 (3.66) | 2.65 (2.49) | 0.72 [0.22, 1.20] | **0.0232\*** |
| 20 min | 4.87 (3.00) | 3.04 (2.40) | 0.77 [0.28, 1.26] | **0.0067\*** |
| 25 min | 5.72 (4.20) | 3.25 (2.31) | 1.07 [0.56, 1.57] | **0.0046\*** |
| 30 min | 5.76 (4.76) | 3.28 (2.20) | 1.13 [0.62, 1.64] | **0.0093** |
| Recovery | 1.93 (1.65) | 1.24 (0.91) | 0.76 [0.27, 1.25] | **0.0384** |

ES: Effect Size (Glass's $\Delta$ ). 95%CI: Confidence Interval 95%. p: P-value from Welch's test. min: minutes. The value for "Baseline" was measured at 0 minutes and "Recovery" at 35 minutes. Glass's $\Delta$ was calculated with the patient group as the reference group.
\*Statistically significant at $p < 0.05$ level with Benjamini-Hochberg correction
\*\*Statistically significant at $p < 0.01$ level with Benjamini-Hochberg correction.

However, in cerebral hemodynamics, a clear difference between the groups was observed. The CBF values were significantly lower in patients (p<0.0001) compared to the healthy group, from the baseline stage to the recovery stage, with a large effect size and negative values, meaning the mean in the patient group was much lower than the healthy ones. Conversely, the values of CVR were significantly higher (p<0.0001) in patients compared to the healthy group, with a large effect size.

The sympathetic-vagal balance indicates a greater sympathetic component in both groups, with similar values from the baseline stage to the first 10 minutes of the orthostatic challenge. From minute 15 onwards, the values in the patients significantly increased as time progressed in the orthostatic stage and remained so in the recovery stage. A change was observed in the effect size, moving from medium at minutes 15 and 20 to a large effect at minutes 25 to 30.

## Discussion

The main findings of this study, beyond the weight difference between groups, include the significant decrease in CBF secondary to the increase in cerebral vascular resistance (CVR) in patients compared to the control group from the supine position. Meanwhile, there were no differences in cardiovascular response between the groups. The shift towards sympathetic dominance was similar between groups from the supine position until the 10th minute of the orthostatic stage, where there was a significant increase in patients compared to controls.

The patients in this study had a history of syncope and a lower BMI compared to the healthy group, and both groups were within normal weight ranges. Greater incidence of syncope related to lower BMI has been reported; Luis Luz Leira et al. (15) in a study of 419 patients undergoing head-up tilt test for syncope investigation reported a higher incidence of a positive test in those with BMI values < 18.5 kg/m2 (p=0.01). Other studies indicate that body weight influences the response to orthostatism, which could be a factor influencing both the history of syncope and the symptomatic response during the test.

Regarding the cardiovascular response in patients and healthy, in the transition from supine to orthostatic position, where gravity causes a redistribution of blood from the upper to the lower body leading to a transient reduction in venous return, cardiac filling, stroke volume, and BP, there was an increase in HR and BP. This response was sufficient to maintain mean arterial pressure at similar values in both groups (1). It is known that the mechanism explaining syncope is the reduction in BP leading to cerebral hypoperfusion (2,3), and this did not occur in the groups studied. On the other hand, a reduction in blood flow can induce a decrease in oxygen or glucose in the brain, triggering symptoms like dizziness, nausea, and weakness, as experienced by patients in this study (17). Therefore, the presence of constant cerebral vasoconstriction during all stages of the test in patients reveals that some of the mechanisms involved in the regulation of cerebral vessels are altered in these patients.

CBF regulation is extremely complex, and the mechanisms involved overlap to ensure adequate oxygen and nutrient supply to the brain. We will discuss two of the main mechanisms involved in regulation: cerebral autoregulation and cerebrovascular reactivity.

### Cerebral Autoregulation

Cerebral autoregulation (CA) is the physiological relationship between CBF and MAP and is defined as the ability of the cerebral vasculature, through changes in cerebral vascular resistance by contracting or relaxing cerebral arteries, to make reflex adjustments in response to BP changes and maintain a constant CBF (17,18). There is evidence that the brain defends itself more effectively against acute hypertension than hypotension (19). Our results indicate that there were no differences in MAP between groups, so it is reasonable to assume that this is not the affected mechanism in the patients studied.

### Cerebrovascular Reactivity

Cerebrovascular reactivity (CVR) is the ability of cerebral vessels to dilate or contract in response to vasoactive substances, increasing or decreasing CBF. Particularly, carbon dioxide (CO2) is a potent vasoactive agent; an increase in partial pressure of CO2 (Pp CO2) causes vasodilation and an increase in CBF by approximately 3% (20), while a reduction in PpCO2 leads to vasoconstriction, making CVR fundamental for cerebral homeostasis (21).

It is known that hyperventilation causes hypocapnia, resulting in cerebral vasoconstriction and a decrease in CBF. A significant limitation in our study is not having measured expired CO2 during the test; however, hyperventilation was observed in 18% of patients during the orthostatic challenge stage, explaining the decrease in CBF during this stage. But it should be noted that this decrease was observed from the supine position and remained throughout the test. Hence, it is necessary to consider other factors influencing this response.

In patients with depression, a decrease in CBF (22,23) has been observed, and in these patients, CVR is reported to be affected or diminished (24,26). The mechanisms affecting cerebral vasomotor function and impacting CBF are diverse, ranging from endothelial response (27), microglia (28), cytokines (29), among others. Anxiety also induces changes in CBF (30,31), particularly due to changes in breathing, with higher levels of anxiety associated with lower CBF during hypocapnia (32). In summary, the intricacies in the mechanisms impacting cerebral vasomotor function and CBF underscore the need to consider a range of both biological and psychological factors when assessing cerebrovascular reactivity. The findings highlight that, in addition to physiological responses to carbon dioxide levels, psychological conditions such as depression and anxiety play a significant role in modulating CBF. This interplay between physical and emotional factors emphasizes the necessity of an integrated approach in the research and management of cerebrovascular disorders.

### Sympathovagal Balance

In the autonomic nervous system response during the test, in the sympathovagal balance, a greater sympathetic component was observed in both groups, with similar values in the first 10 minutes, and after this time, a significantly greater increase was observed in patients compared to the healthy group. Although there are abundant sympathetic nerve fibers in the cerebral vasculature, their influence on CBF is not yet fully understood. However, some studies indicate that sympathetic activation can affect CBF (33,34), suggesting that the effect of increased sympathetic activity in patients contributes to maintaining adequate cardiovascular response during the orthostatic challenge and adds to the factors preserving the decrease in CBF in these patients.

In summary, our study provides insight into the reduction of CBF in young and healthy individuals with a history of syncope or pre-syncope from the supine position, which may be due to the combination of various factors and exacerbated in the standing position, and might also include genetic susceptibility in some (35, 36). However, there is much to explore to better understand the factors that may influence this reduction. Further study of this topic could help identify effective prevention and treatment strategies for this group of individuals. Therefore, it is clear that future research in this area is not only valuable but also necessary.

## Data Availability

All data produced in the present study are available upon reasonable request to the authors

## Acknowledgments

We thank the nursing team for their help during the test and to Dr. Andrei Kostine, for his valuable support in obtaining the measurement of cerebral blood flow

## Sources of funding

No specific financial support was provided by any agencies in the public, commercial, or not-for-profit sectors for the conduct of this research.

## Disclosures

Nothing to disclose

## Notes

### Competing Interest Statement

The authors have declared no competing interest.

### Funding Statement

This study did not receive any funding

### Author Declarations

Ethics and Research Committees of the National Institute of Cardiology Ignacio Chavez gave ethical approval (14/904) for this work.

